# Systemic immune markers and infection risk in preterm infants fed human milk fortified with bovine colostrum or conventional fortifier, a secondary analysis of the *FortiColos* trial

**DOI:** 10.1101/2023.11.01.23297894

**Authors:** Ole Bæk, Tik Muk, Lise Aunsholt, Gitte Zachariassen, Per Torp Sangild, Duc Ninh Nguyen

## Abstract

**Background:** For very preterm infants, human milk is often fortified with formula products based on processed bovine milk. Intact bovine colostrum (BC) is rich in anti-inflammatory milk factors and considered an alternative. Our objective was to investigate if BC affects anti-inflammatory/T_H_2 immunity and infection risk in very preterm infants.

**Methods:** In a secondary analysis of a multicenter, randomized controlled trial (NCT03537365), very preterm infants (26-31 weeks gestation, 23% small for gestational age, SGA) were randomized to receive BC (ColoDan, Biofiber, Denmark, n=113) or a conventional fortifier (PreNAN, Nestlé, Switzerland, n=116). Infection was defined as antibiotic treatment for five or more consecutive days. Levels of 29 cytokines and chemokines were measured in plasma before and after start of fortification.

**Results:** Infants fortified with BC showed more infection episodes (20 vs. 12%, P<0.05) and tendency to higher cumulative infection risk (hazard ratio, HR 1.9, P=0.06), particularly for SGA infants (HR 3.6, P<0.05). Additionally, BC-fortified infants had higher levels of T_H_2 related cytokines and chemokines (IL-10, MDC, MCP4) and reduced levels of cytokines related to T_H_1/T_H_17 responses (IL-15, IL-17, GM-CSF). The differences were most pronounced in SGA infants, displaying higher levels of T_H_2-related IL-4, IL-6, and IL-13, and lower interferon-γ and IL-1α levels in the BC group

**Conclusion:** Infants fortified with BC show delayed transition from T_H_2-to T_H_1-biased systemic immunity, especially for SGA infants. This was associated with more frequent antibiotic use, indicating elevated sensitivity to infection. Thus, an anti-inflammatory milk supplement like BC may delay systemic immune development in preterm infants with effects depending on weight at birth.

## Introduction

Very preterm infants (born before 32 weeks of gestation) show greater risk of infections in early life, with risks increasing with the degree of immaturity, particularly in those born small for gestational age (SGA) (1,2). These vulnerable infants are also at risk of post-natal growth failure, partly due to co-morbidities and systemic infections, but also because enteral feeding poses challenges, with risks of feeding intolerance, metabolic dysfunction and necrotizing enterocolitis (NEC) (3). Mother’s own milk (MOM) is the preferred milk diet for infants born preterm and reduces the risk of early life infections compared with infant formula (4–6). MOM contains a multitude of bioactive compounds such as antimicrobial peptides, immunomodulatory proteins, commensal microorganisms, maternal leucocytes and prebiotics (7). Unfortunately, MOM is often unavailable, or not sufficient in amounts or nutritional composition sufficient to nourish infants for optimal growth after preterm birth. In many countries, donor human milk (DHM) is used as a substitute for MOM. However, DHM is obtained from mothers in late lactation, with relatively low level of nutritional and bioactive proteins, partly degraded by heat pasteurization (8). Adequate growth can be attained when MOM and DHM are fortified with nutrient fortifiers, often based on bovine milk formula products (9). Such fortifiers are highly processed and proteins often pre-hydrolyzed to enhance amino acid absorption and avoid constipation and allergic reactions (10).

Recently, intact bovine colostrum (BC) has been proposed as an alternative nutrient fortifier for preterm infants (11,12). Like human colostrum, BC is rich in protein and milk bioactive and immune-modulating compounds, like immunoglobulin G and A, lactoferrin, multiple growth factors (13–16). When gently dried, heat-pasteurized and irradiated to obtain a near-sterile product suitable for clinical use, bioactivity of BC proteins is largely preserved (17,18). Based on our previous pre-clinical research on BC in preterm pigs (19–23) and reports from clinical trials in preterm infants (12,24,25), BC fortification may modulate immune development and infection risk, potentially by promoting a more anti-inflammatory immune phenotype.

A recent randomized controlled trial in very preterm infants (n=232) investigated if BC fortification could adequately support infant growth during hospitalization, compared with a conventional fortifier (CF) (26). The results showed similar body growth between groups, and BC fortification improved bowel habits, as indicated by less use of laxatives (26,27). However, in the per-protocol analysis there was a tendency to more use of antibiotics in the BC group. In this secondary analysis of the data, we explore in detail the impact of BC fortification on infection risk and development of systemic immunity, based on levels of cytokines and chemokines in plasma collected before and after start of fortification. When adjusting for relevant confounders, we confirm that BC fortification is associated with increased risk of infection, especially in SGA infants, and with increased levels of anti-inflammatory cytokines.

## Methods

### Study design

The registered protocol (clinicaltrials.gov: NCT03537365) has been published along with the main clinical finding (26,28). Briefly, very preterm infants (26-31 weeks of gestation, n=232, 23% SGA), from eight neonatal units in two regions of Denmark, were randomized to receive either BC (pasteurized, spray-dried and irradiated powder, Biofiber, Denmark) or a conventional bovine-based fortifier (CF, FM85 PreNAN, Nestlé, Switzerland). All infants received an enteral diet of MOM and/or DHM and fortification was initiated when enteral feeding volumes reached 100-140 mL/kg/day and blood urea nitrogen was <5 mmol/L. Fortification was continued until a gestational age of 34+6 weeks or discharge. The amount of fortification followed local guidelines and international recommendations, with added protein not exceeding 1.4 g per 100 mL of human milk (29). Infants with major congenital malformations, gastrointestinal surgery or receiving infant formula before start of fortification were excluded. Written informed consent was obtained from all participants’ parents or guardians. In cases of withdrawn informed consent, data collected up to that point in time was used, if allowed by parents/guardians. Infants were classified as SGA by a birthweight Z-score less than two standard deviations for their gestational age.

### Antibiotics use and infection incidence

Use of all medications were registered during the trial, both before and after start of fortification. In the current analysis, the type and duration of intravenous antibiotic medications were identified and calculated. In three cases (1 CF/SGA and 2 BC/AGA), records on prescribed medications were unavailable and these infants were removed from further analyses. A list of the prescribed antibiotics is shown in Supplementary Table S1.

An episode of infection was pre-defined in the study protocol as five or more consecutive days on any type of intravenous antibiotics, regardless of blood culture or other biochemical findings (28). As such, early-onset infection was defined as five or more consecutive days of antibiotics, starting within the first 48 hours after birth, while late-onset infections would occur after 72 hours post-partum. Two periods of antibiotic use more than 24 hours apart were considered as inconsecutive. Clinical blood samples, measuring C-reactive protein (CRP), blood gas and bacterial cultures, were not collected consistently across the units in relation to suspected infections. Hence, these confirmatory blood results were available only from a subset of the infants with suspected infection.

### Blood sampling and cytokine measurements

Blood samples were collected before fortification and at approximately one (7±1 days) and two weeks (14±2 days) after start. Blood was collected by capillary puncture in EDTA-coated tubes, cooled down and centrifuged (2500 x g, 4°C, 10 minutes) for plasma collection within 4 hours and stored at -20°C until further analysis. Levels of 29 plasma pro- and anti-inflammatory cytokines and chemokines related to T_H_1/T_H_2/T_H_17 polarization were measured by multiplex, fluorescent immunoassays as per the manufacturer’s instructions (V-plex proinflammatory cytokine/chemokine analysis panels, Meso Scale Diagnostics, USA). Levels of markers below the detection limits, but higher than blank values, were set to half of the lowest standard for each individual cytokine or chemokine.

### Statistics

All statistics were performed using Stata 14 (StataCorp, USA). Before the start of fortification, group differences in antibiotic use and number of infectious episodes were calculated using a Chi-square or student t-test, depending on data type. After the start of fortification, the effect of the intervention on incidence of infections were calculated using a logistic regression model while the cumulative risk of infection was calculated with a cox proportional hazard, conditional risk set model, accounting for multiple infections per infant (30). Duration of antibiotic treatment and cytokine levels were evaluated using a generalized linear model. In models, sex, SGA status, gestational age, Apgar score and antibiotics use prior to intervention was used as cofactors. During recruitment it became evident that randomization was unevenly distributed in the Eastern and Western regions of Denmark, and feeding practices and probiotics use also differed. For that reason, geographical region was also added as a covariate (26). Plasma cytokines and chemokines were analyzed by a mixed effect model and assay plate number was used as a random factor to control for any inter-plate differences. If residuals of models could not conform to normality, data was log transformed. Any data that could not conform to normality was analyzed with an appropriate non-parametric model. All models were performed first on all infants, and then stratified according to SGA or AGA status, using the same covariates. Due to the lower number of SGA infants, incidence of infections was calculated with a penalized logistic regression (31) and cumulative infection risk using the log-rank test. P values less than 0.05 were considered statistically significant, while values less than 0.1 were considered as a tendency to a significant effect.

## Results

### Infection risk and antibiotics use before start of fortification

Postnatal age at start of intervention was about one day shorter in the BC group (Table 1, P<0.05), however this difference was not apparent for SGA infants (Table 1). Other baseline characteristics for infants did not differ between diet or AGA/SGA groups, except that Apgar scores were lower in BC-fortified infants (but only in AGA infants, P<0.01, Table 1). Incidences of early- or late-onset infections, before start of the intervention, did not differ between groups, with or without AGA/SGA stratification, although a tendency to fewer late-onset infections was observed in BC-fortified AGA infants (6 vs 13%, P=0.09, Table 1). Before start of intervention, about 50% of all the infants received antibiotics, with similarly distributed between early- and late-onset infections, and between BC and CF groups, with or without SGA (Table 1, all P>0.1). No infant had more than one late-onset infection before start of fortification while two infants had both an early and late onset infection (one from each group). Similarly, the duration of antibiotic treatment before start of intervention, did not differ between groups, with/without AGA/SGA stratification (Table 1, all P>0.1).

**Table 1:** Baseline characteristics and infection risk before start of fortification.

|  | All infants |  |  | AGA |  |  | SGA |  |  |
| --- | --- | --- | --- | --- | --- | --- | --- | --- | --- |
|  | CF<br>n=116 | BC<br>n=113 | P | CF<br>n=91 | BC<br>n=85 | P | CF<br>n=25 | BC<br>n=28 | P |
| Age at fortification, days | 9.4<br>(3.7) | 8.4<br>(2.8) | <0.05 | 9.6<br>(4.1) | 8.5<br>(2.9) | <0.05 | 8.5<br>(2.9) | 8.2<br>(2.7) | NS |
| Gestational age at birth, days | 200<br>(10) | 201<br>(10) | NS | 200<br>(11) | 202<br>(10) | NS | 202<br>(9) | 200<br>(9) | NS |
| Male sex | 54%<br>(63/116) | 60%<br>(68/113) | NS | 52%<br>(47/91) | 59%<br>(50/85) | NS | 64%<br>(16/25) | 64%<br>(18/28) | NS |
| Birthweight, g | 1167<br>(322) | 1175<br>(333) | NS | 1242<br>(309) | 1285<br>(289) | NS | 894<br>(198) | 840<br>(211) | NS |
| Cesarian section | 74%<br>(86/116) | 70%<br>(79/113) | NS | 67%<br>(61/91) | 61%<br>(52/85) | NS | 100%<br>(25/25) | 96%<br>(27/28) | NS |
| **Complications during birth<sup>†</sup>** |  |  |  |  |  |  |  |  |  |
| Preeclampsia | 21%<br>(23/112) | 23%<br>(26/111) | NS | 10%<br>(9/89) | 14%<br>(12/83) | NS | 61%<br>(14/23) | 50%<br>(14/28) | NS |
| Premature rupture of membranes | 21%<br>(24/116) | 20%<br>(23/113) | NS | 24%<br>(22/91) | 26%<br>(22/85) | NS | 8% (2/26) | 4%<br>(1/28) | NS |
| Chorioamnionitis | 3%<br>(3/113) | 4%<br>(5/113) | NS | 2%<br>(2/89) | 6%<br>(5/85) | NS | 4%<br>(1/25) | 0%<br>(0/28) | - |
| Placental abruption | 6%<br>(7/114) | 12%<br>(13/111) | NS | 8%<br>(7/89) | 15%<br>(13/84) | NS | 0%<br>(0/26) | 0%<br>(0/27) | - |
| Mean APGAR score at 5 min | 9.3<br>(±1.4) | 8.7<br>(±1.9) | <0.05 | 9.4<br>(±1.2) | 8.6<br>(±2.1) | <0.01 | 8.8 (±1.8) | 9.3<br>(±0.7) | NS |
| APGAR score <7 at 5 min | 7%<br>(8/116) | 11%<br>(12/113) | NS | 5%<br>(4/91) | 14%<br>(12/85) | 0.03 | 17%<br>(4/25) | 0%<br>(0/28) | - |
| **Infections and antibiotics use prior to intervention** |  |  |  |  |  |  |  |  |  |
| Early onset infection<sup>§</sup> | 10%<br>(12/116) | 12%<br>(13/113) | NS | 9%<br>(8/91) | 8%<br>(7/85) | NS | 15%<br>(4/26) | 21%<br>(6/28) | NS |
| Late onset infection<sup>#</sup> | 13%<br>(15/116) | 8%<br>(9/113) | NS | 13%<br>(12/91) | 6%<br>(5/85) | 0.09 | 12%<br>(3/25) | 14%<br>(4/28) | NS |
| Any AB | 47%<br>(55/116) | 54%<br>(61/113) | NS | 46%<br>(42/91) | 53%<br>(45/85) | NS | 52%<br>(13/25) | 57%<br>(16/28) | NS |
| Total AB use, days | 3.0<br>(±4.2) | 3.4<br>(±5.0) | NS | 3.0<br>(±4.3) | 3.1<br>(±4.9) | NS | 3.4<br>(±4.0) | 4.4<br>(±5.4) | NS |
| AB use during early onset infections, days | 8.8<br>(±1.4) | 11.2<br>(±1.7) | NS | 8.8<br>(±2.0) | 10.7<br>(±2.8) | NS | 9.0<br>(±1.3) | 11.7<br>(±2.2) | NS |
| AB use during late onset infections, days | 10.8<br>(±1.0) | 13.8<br>(±1.5) | NS | 11.3<br>(±1.3) | 15.2<br>(±2.3) | NS | 9.0<br>(±1.0) | 12.0<br>(±2.1) | NS |

Baseline characteristics, risk of infections and use of intravenous antibiotics (AB) in very preterm infants randomized to be fortified with either conventional fortifier (CF) or bovine colostrum (BC). Shown for all infants and stratified for birth weight status. AGA: Birthweight appropriate for gestational age, SGA: Small for gestational age, NS: Not significant. †: Information on birth complications could not be obtained for all infants, see individual n numbers. §: Defined as more than 5 consecutive days on AB, starting within 48 hours of birth. #: Defined as more than 5 consecutive days on antibiotics, starting more than 48 hours of birth. Binomial data shown as percentage and continuous data shown as means with corresponding standard deviation.

### Infection risk and antibiotic use after start of fortification

Following the start of fortification, 52 episodes of infection were observed across 37 infants. Results of clinical blood cultures or samples collected around the time of antibiotic treatment are shown in Supplementary Table 2. Blood cultures were only performed in 50% (26/52) of reported infection episodes and 65% (17/26) of these showed growth of a possibly pathogenic bacteria, with no significant differences in the culture positive rate or pathogen found between the groups. Likewise in available samples CRP, pH and lactate levels were similar between groups. Of the 52 reported episodes of infection, in 42 (14 CF, 28 BC) CRP was >10 ug/mL, with/without positive blood culture, while in four episodes (2 CF, 2 BC) CRP was <10 ug/mL and blood cultures negative/not performed. In six cases (4 CF, 2 BC),) no CRP or blood culture results were available.

The overall risk of infection following fortification was most strongly associated with gestational age (P<0.001, Supplementary Figure S1A), but not with postnatal age at start of fortification (P>0.1, Supplementary Figure S1B). Likewise, SGA status or antibiotic use before start of fortification was associated with later infection (both P<0.05, Supplementary Figure S1 CS1C,D).

Across all infants, the incidence of at least one infectious episode was higher in the BC group (Table 2, 20 vs. 12%, P<0.05). In SGA infants, the mean incidence of infection was twice as high in BC-fortified SGA infants vs. CF-fortified SGA infants, but not significantly (Table 2, 32 vs. 15%, P>0.1). There was a tendency towards higher cumulative infection risk among all BC-fortified infants (Cox model HR: 1.9, CI95%: 1.0-3.7, P=0.06, Figure 1A), with no difference in risk among AGA infants (HR: 1.4, CI95%: 0.6-3.3, P>0.1, Figure 1B). However, cumulative infection risk among SGA infants was significantly higher in those receiving BC fortification (risk ratio, RR: 3.3, CI95%: 1.1-10.2, P<0.05, Figure 1C). In a sensitivity analysis, including only the 42 cases of infection with increased CRP and/or positive blood culture, infection risk was significantly increased across all BC-fortified infants (HR: 2.3, CI95%: 1.1-4.3, P<0.05, Supplemental Figure S2A), as well as for BC-fortified AGA infants (HR: 2.3, CI95%: 1.0-5.4, P=0.06, Supplemental Figure S2B) and SGA infants (RR: 3.0, CI95%: 1.0-9.2, P<0.05, Supplemental Figure S2C)

**Figure 1:**
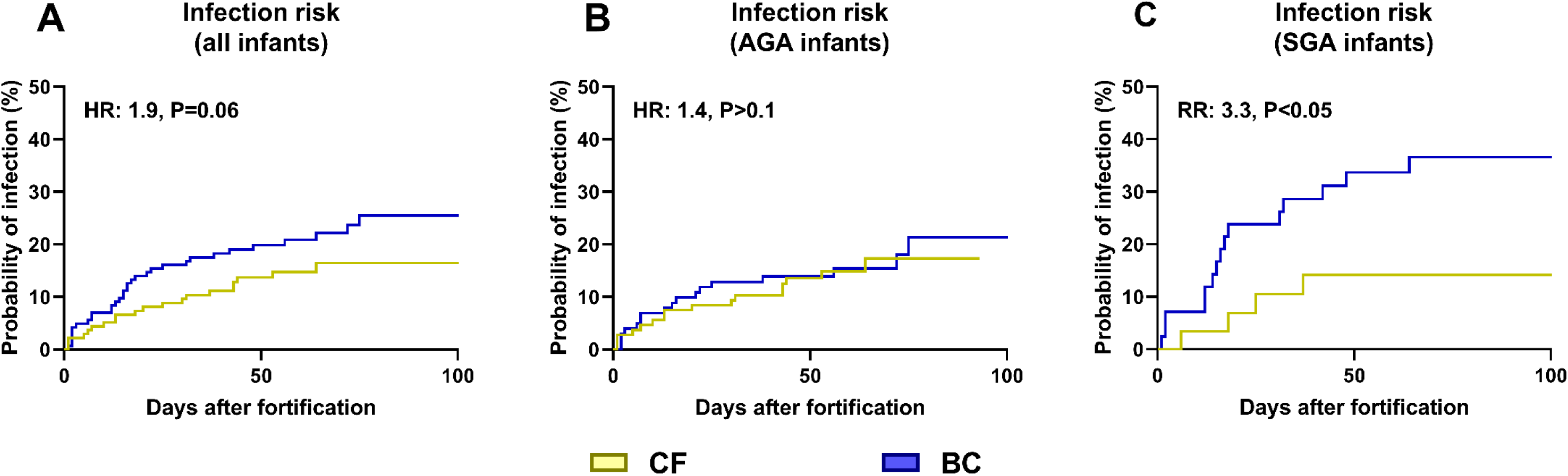
Time to infectious episodes in infants fortified with bovine colostrum (BC) or conventional fortifier (CF), shown as Kaplan Meyer curves with results of corresponding Cox proportional hazard models or log rank test shown as text. Shown for all infants (**A**) or stratified by birth weight status (**B**: AGA, appropriate for gestational age; **C:** SGA, small for gestational age). HR: Hazard ratio, RR: Risk ratio.

**Table 2:** Risk of infections and use of antibiotics and parenteral nutrition after start of fortification.

|  | All infants |  |  | AGA |  |  | SGA |  |  |
| --- | --- | --- | --- | --- | --- | --- | --- | --- | --- |
|  | CF<br>n=116 | BC<br>n=113 | P | CF<br>n=91 | BC<br>n=85 | P | CF<br>n=25 | BC<br>n=28 | P |
| **Infections<sup>#</sup>** |  |  |  |  |  |  |  |  |  |
| Incidence of infection | 12%<br>(14/116) | 20%<br>(23/113) | 0.04 | 10 %<br>(10/91) | 16%<br>(14/85) | NS | 16 %<br>(4/25) | 32 %<br>(9/28) | NS |
| Multiple infections | 29%<br>(4/14) | 26%<br>(6/23) | NS | 40%<br>(4/10) | 14%<br>(2/14) | NS | 0%<br>(0/4) | 44%<br>(4/9) | - |
| AB without infection<sup>†</sup> | 9%<br>(10/116) | 7%<br>(8/113) | NS | 10%<br>(9/91) | 6%<br>(5/85) | NS | 4 %<br>(1/25) | 10%<br>(3/28) | NS |
| **Antibiotics** |  |  |  |  |  |  |  |  |  |
| Total use, days | 1.9<br>(±5.4) | 3.6<br>(±9.5) | 0.02 | 2.1<br>(±5.9) | 2.9<br>(±1.0) | NS | 1.4<br>(±3.0) | 5.5<br>(±8.5) | 0.06 |
| Without infection, days | 0.3<br>(±0.8) | 0.3<br>(±1.1) | NS | 0.3<br>(±0.1) | 0.2<br>(±0.1) | NS | 0.1<br>(±0.7) | 0.7<br>(±1.7) | NS |
| With infection, days | 13.9<br>(±8.9) | 16.2<br>(±15.5) | NS | 16.3<br>(±9.5) | 16.5<br>(±19.2) | NS | 8.0<br>(±1.4) | 15.8<br>(±8.0) | NS |
| **Parenteral nutrition** |  |  |  |  |  |  |  |  |  |
| Incidence | 28%<br>(32/116) | 31%<br>(35/113) | NS | 23%<br>(21/91) | 26%<br>(22/85) | NS | 44%<br>(11/25) | 46%<br>(13/28) | NS |
| Duration, days | 1.5<br>(±1.5) | 1.6<br>(±1.5) | NS | 1.6<br>(±1.7) | 1.7<br>(±1.2) | NS | 1.4<br>(±1.3) | 1.4<br>(±1.8) | NS |

Risk of infections and use of intravenous antibiotics (AB) and parenteral nutrition in very preterm infants fortified with either conventional fortifier (CF) or bovine colostrum (BC). Shown for all infants and stratified for birth weight status. AGA: Birthweight appropriate-for-gestational age, SGA: Small-for-gestational age, NS: Not significant. #: Infection defined as 5 or more consecutive days on antibiotics after start of fortification.†: AB prescribed for less than 5 days after start of fortification. Binomial data shown as percentage and continuous data shown as means with corresponding standard deviation.

Across all infants, total length of antibiotic treatment was longest in the BC group, although this is explained by the higher incidence of infection as length of antibiotics treatment did not differ among infants without infection (Table 2). Again, the effects were most pronounced for the SGA subgroups (Table 2). Importantly, the use of parenteral nutrition, an additional risk factor for postnatal infection, did not differ after start of fortification (Table 2). Interestingly, there was interaction between use of antibiotics prior to start of fortification (for any duration of time), and later risk of infection. Without antibiotics use, infants fortified with CF showed lower infection risk later than the corresponding BC infants, or infants receiving antibiotics before start of fortification (Supplementary Figure S3 A-C).

### Plasma cytokines before and after start of fortification

Before fortification no differences in plasma cytokines were observed (Figure 2A-L). One week after start of the intervention, BC infants had significantly higher levels of T_H_2 cytokine IL-10 (Figure 2A, P<0.01, both AGA and SGA) with a tendency to higher IL-4 levels (Figure 2B, P=0.07 for SGA infants). At the same time, BC-fortified infants had lower levels of IL-5, IL-15, IL-17 and granulocyte-macrophage colony-stimulating factor (GM-CSF, Figure 2C-F, all P<0.05). BC-fortified SGA infants also tended to have lower levels of interferon-γ (IFN-γ, Figure 2G, P=0.07). The above differences between diet were less pronounced at two weeks after start of fortification. However, differences persisted within the SGA subgroup for IL-10, IL-15 and GM-CSF (Figure 2A,C,E). In addition, levels of IL-6 and IL-13 were increased for BC-fortified SGA infants (Figure 2H, I, both P<0.01), while the levels of GM-CSF and IL-1α were decreased (Figure 2J, P<0.01). Interestingly, BC-supplemented AGA infants showed lower IL-6 levels both one and two weeks after start of fortification (Figure 2H, P<0.01), contrasting the above effects in SGA infants.

**Figure 2:**
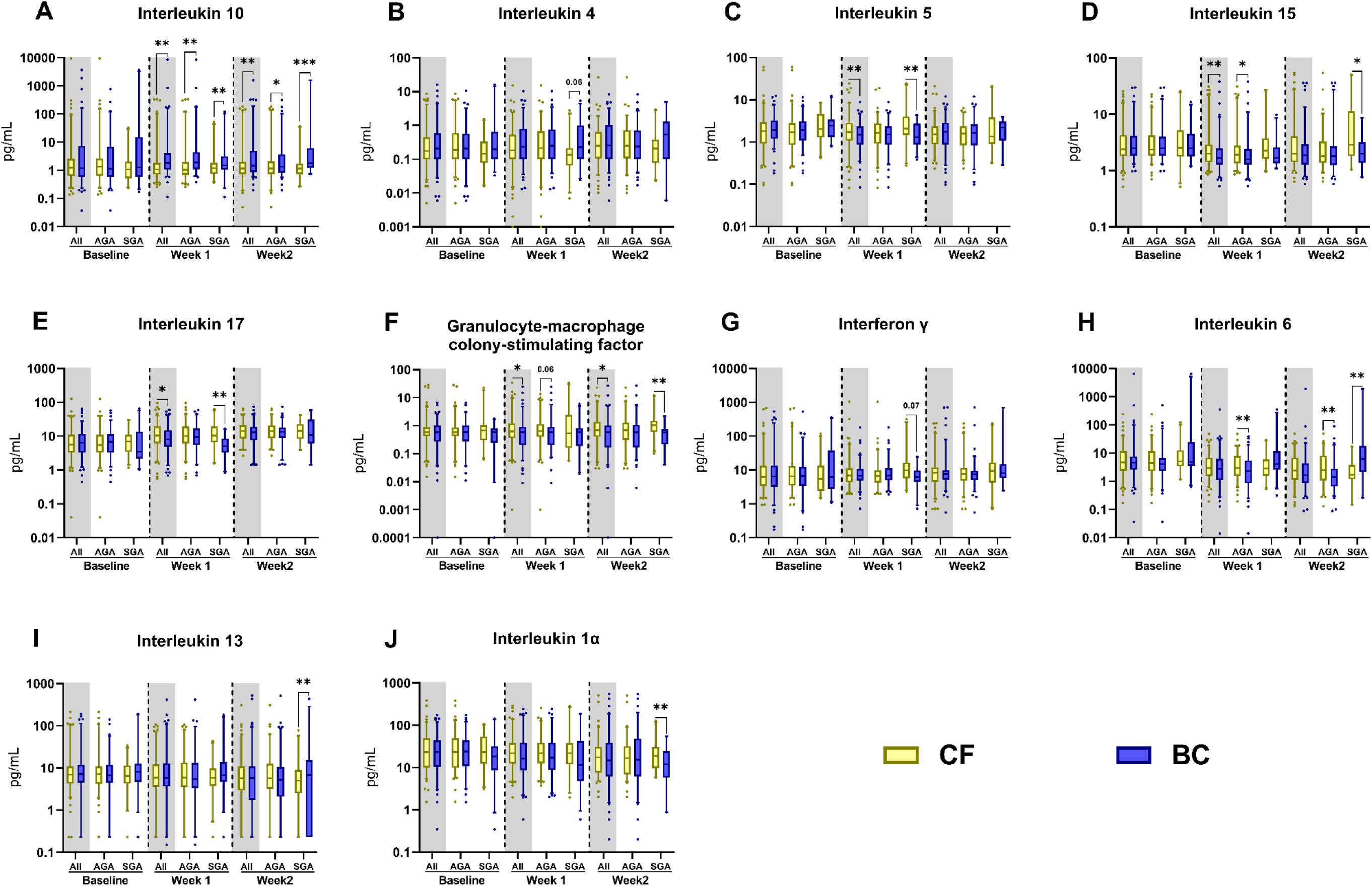
Plasma cytokine levels in infants fortified with bovine colostrum (BC) or conventional fortifier (CF). Shown before start of fortification (Baseline) and one and two weeks after start of fortification for all infants or stratified by birth weight status (AGA, appropriate for gestational age; SGA, small for gestational age). Shown as 95% percentile box plots for differences between BC and CF fortification, *: P<0.05, **: P< 0.01, while P values between 0.05 and 0.1 are shown as text.

To investigate whether cytokine levels and their responses to fortification were influenced by the presence or absence of suspected infection, a sensitivity analysis, with removal of the 37 infants with infection, was conducted. In this analysis, the majority of the previously reported differences persisted. Specifically, SGA infants fed with BC continued to show higher levels of IL-6, IL-10, and IL-13 (all with P<0.05), along with decreased levels of GM-CSF, IL-1α, IL-1β, IL-5, and IL-17 (all with P<0.05) at one and/or two weeks after the initiation of fortification (data presented in Supplementary Table S3).

### Plasma chemokines before and after start of fortification

The effects of BC fortification on circulating chemokine levels were less obvious. BC-fortified infants showed higher levels of monocyte chemoattractant protein 4 (MCP-4 or CCL13) and IFN-γ inducible protein-10 (IP-10 or CXCL10) one and two weeks after the start of fortification. This was driven mainly by effects among AGA infants (Figure 3A,B, P<0.01 and P<0.05), with a similar trend observed for thymus and activation regulated chemokine (TARC or CCL17, Figure 3C, P=0.06 and P=0.07).

**Figure 3:**
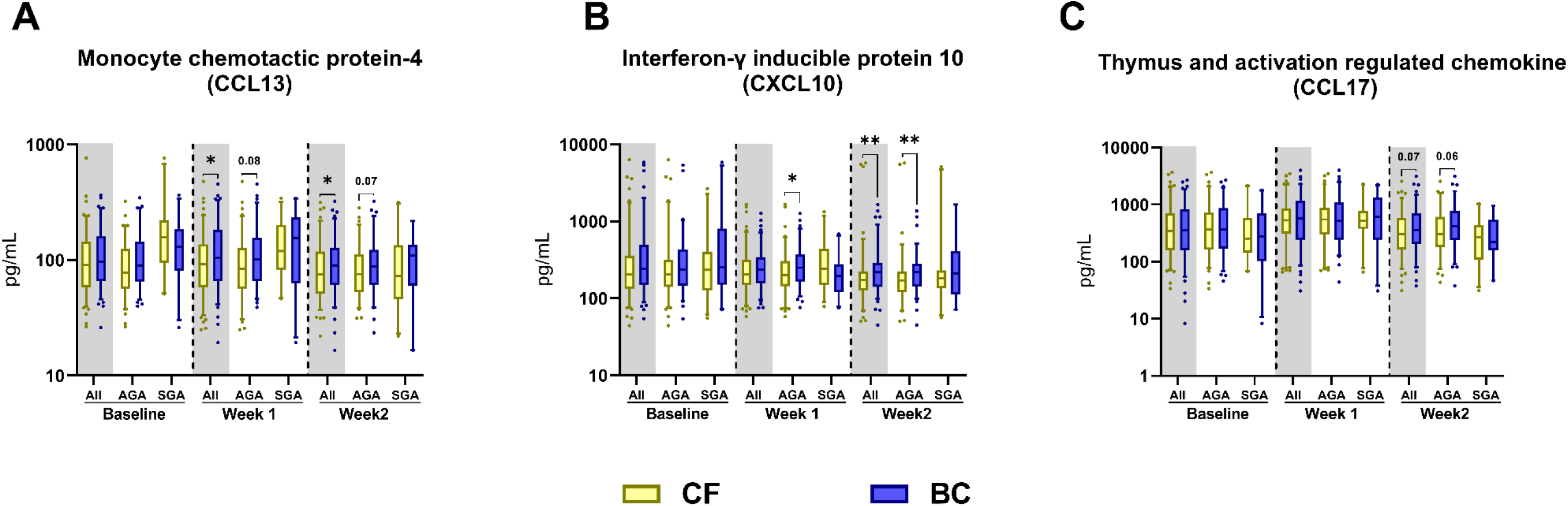
Chemokine profiles in infants fortified with bovine colostrum (BC) or conventional fortifier (CF). Shown before start of fortification (Baseline) and one and two weeks after start of fortification for all infants and stratified by birth weight status (AGA, appropriate for gestational age; SGA, small for gestational age). Shown as 95% percentile box plots for differences between BC and CF fortification, *: P<0.05, while P values between 0.05 and 0.1 are shown as text.

## Discussion

Securing adequate enteral nutrition in preterm infants is key to achieve optimal postnatal growth but the choice of milk diet may also influence other aspects of health, including systemic immunity. We found that the use of BC as a fortifier to human milk (MOM, DHM or a mixture) was associated with an increased use of intravenous antibiotics, indicating an increased incidence of infections, especially in infants born SGA, even after excluding infants without positive blood cultures or high CRP levels. Fortification with BC was also associated with a blood immune profile that reflected higher levels of anti-inflammatory/T_H_2/T_Reg_ cytokines (e.g IL-10) and lower levels of pro-inflammatory/T_H_1/T_H_17-related cytokines (e.g. IL-15, IL-17, GM-CSF), most clearly after one week of fortification. Again, this pattern was most clear for SGA infants, where infants fortified with BC also showed higher levels of T_H_2 cytokines (IL-4, IL-6, IL-13) and a tendency to lower levels of key T_H_1 cytokines (IFN-γ, IL-1α). These differences therefore indicate that the immune profile of BC-fortified infants, especially those born SGA, could be skewed towards a more anti-inflammatory/T_H_2 driven immune phenotype. Importantly, these effects persisted after excluding infants that experienced an infectious episode after start of fortification, suggesting that the observed effects were driven by the diet intervention and not by inflammation associated with infections. However, IL-5, a classical T_H_2 cytokine related to eosinophil activation showed the opposite effect with lower levels in BC-fortified infants. This could be explained by T_Reg_ derived IL-10 which is known to suppress IL-5 production at mucosal surfaces (32,33). Together with the observed interaction of BC effects on infection risk with antibiotics use prior to the start of fortification, this indicates that BC effects are mediated via the gut mucosal immune system and/or the gut microbiota. We are currently investigating stool samples collected during the trial to elucidate any effects BC fortification and antibiotics had on gut microbial composition.

The increased infection risk in BC infants was unexpected, as previous use of BC supplementation in humans and animals indicates protection against gut and respiratory infections (11,34–38). However, these results may not be valid for supplementation of BC to a very preterm infant, with an immature gastrointestinal tract and immune system. In preterm pigs, exclusive or partial BC feeding just after birth promotes gut maturation, prevents NEC, improves bacterial clearance during systemic infections and increases the number of circulating T_Reg_ cells, suggesting an impact on systemic immunity (20,22,39,40). Likewise, administration of BC to adult mice reduced their blood immune cell pro-inflammatory responses (41,42). Previously, three larger randomized and one pilot trial of BC supplementation to preterm infants have been conducted (12,24,25,43). In one of these trials, administration of BC just after preterm birth, with no MOM feeding, reduced the incidence of severe late-onset sepsis, defined as infection-related organ dysfunction. However, BC had no impact on overall infection risk or antibiotic use but was linked to increased blood T_Reg_ cells in the weeks following preterm birth (27). Another recent study, supplementing MOM with BC instead of formula in the first week after birth, showed no effects on systemic infections. However extensive use of antibiotics immediately following birth in the trial may have clouded any BC effects (26).

In our study, we identified infections based on antibiotics use and could not judge disease severity, but increased IL-10 levels in BC infants may result from more circulating T_Reg_ cells. The resulting anti-inflammatory/T_H_2/T_Reg_-biased immunity may predispose to systemic infections later. For instance, very preterm infants diagnosed with severe systemic infections show reduced capacity to mount a pro-inflammatory responses prior to infection onset (44). However, BC may lead to reduced pro-inflammatory responses, with an impaired ability to fight infections, depending on other variables, such as birth status (SGA/AGA), postnatal age and milk diet (DHM, MOM). Possibly, anti-inflammatory effects of BC supplementation also affect preterm infants differently compared with term infants or older children. Compared with term infants, preterm infants already show diminished pro-inflammatory responses at birth, while their ability to produce IL-10 is less affected or even improved (44–47). This immature immune phenotype is further affected by fetal growth restriction, induced by a variety of antenatal factors, as cord blood immune cells from SGA and intrauterine growth restricted preterm infants show an even lower capacity to produce pro-inflammatory cytokines (48–50). It is therefore plausible that BC increase the infection susceptibility specifically in SGA infants because they already show low pro-inflammatory capacity. In our study, the infection risk was similar in CF- and BC-fortified AGA infants, even if they showed differences in plasma cytokine levels. Finally, it cannot be excluded that anti-inflammatory BC effects negatively affect systemic infections in early life of preterm infants, but positively affect immune responses at mucosal surfaces (gut, lungs, skin) (35,36).

Our study has several limitations. Given the secondary nature of the analysis, we cannot define the causal links between plasma cytokine profiles and infection risk. Despite randomization there were uneven distributions in some factors between the BC and CF groups, including region of birth and infection risk prior to start of fortification. Although we included these factors as covariates in our statistical models, residual confounding cannot be ruled out. It is also important to note that our trial was not designed and powered to investigate effects of BC on risk of infections but had a focus on safety and feasibility to secure adequate growth rates (26). Consequently, only a part of the infants with suspected infections had blood sampled to for bacterial culture and to assess CRP levels and since the trial was not blinded, we cannot exclude possible bias of clinicians. Yet, we find it unlikely that clinicians would be more likely to prescribe antibiotics to BC-fortified infants. In all, our findings do raise important questions regarding the influence of continued feeding with a supplementary milk diet like BC, favoring anti-inflammatory immune responses. Especially in already immune-suppressed SGA infants, this may pose a risk, at least for systemic infections. Larger studies, controlling for age and weight at birth, milk diet and antibiotics treatment prior to intervention are required to elucidate these highly complex and possibly interacting effects.

## Data Availability

All data produced in the present study are available upon reasonable request to the authors

## Ethical considerations

This study adhered to the principles outlined in the Declaration of Helsinki and was conducted in accordance with the ethical standards of the Danish National Center for Ethics. The trial was approved by the Scientific Ethical Committee of the Region of Southern Denmark (S-20170095) and the Danish Data Protection Agency (17/33672). An independent data safety monitoring board reviewed trial data and safety during the enrolment period, incorporating preliminary assessment of key outcomes and potential adverse events. Written informed consent was obtained from all participants’ parents or guardians before enrollment in the study.

## Author contributions

GZ, LA and PTS planned and executed the original trial. The secondary analysis was planned by OB and DNN. Laboratory work and data analysis was performed by OB and TM. OB wrote the first draft of the manuscript while PTS and DNN held joint responsibility for the final contents and all authors read and approved the final manuscript.

