## Supplementary Figures and Tables for "Systemic immune markers and infection risk in preterm infants fed human milk fortified with bovine colostrum or conventional fortifier, a secondary analysis of the *FortiColos* trial"

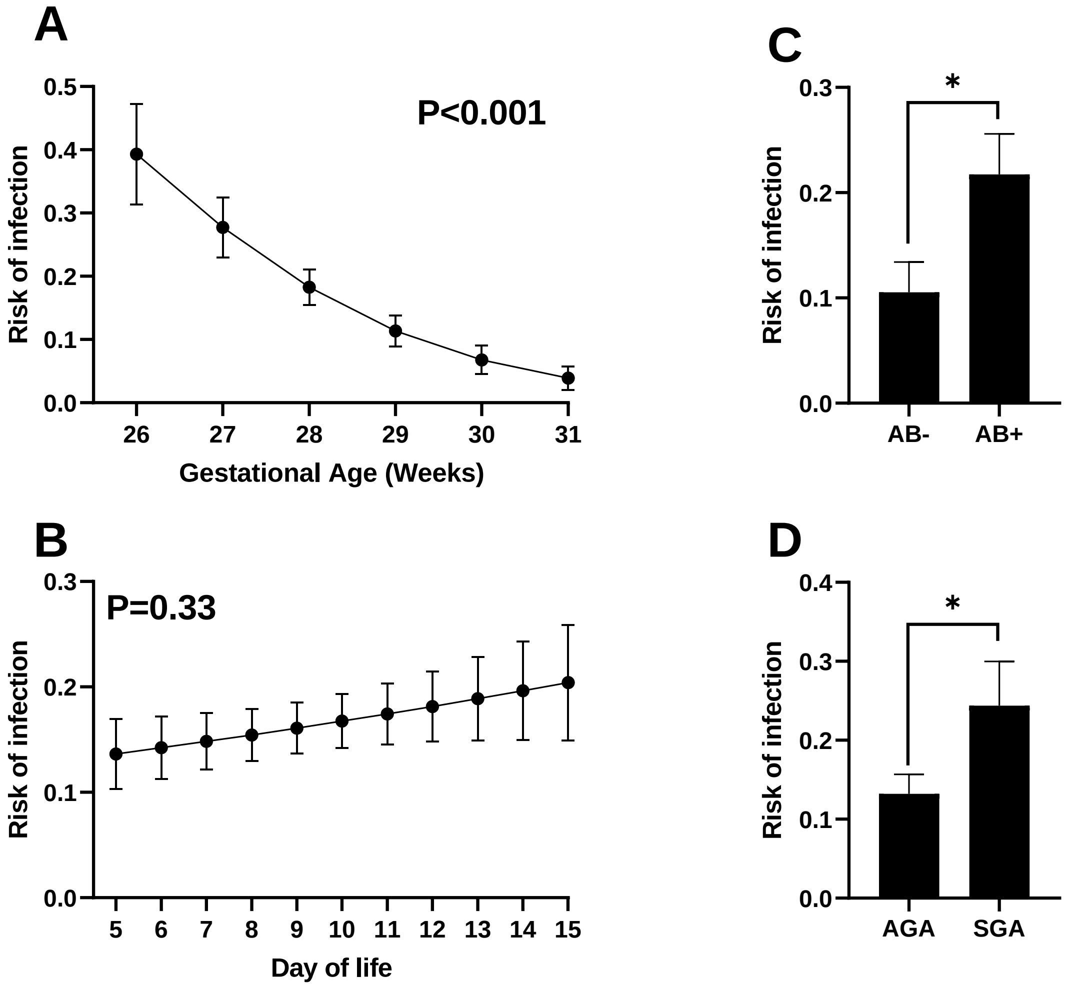


**Supplementary Figure S1**: Relationship between gestational age at birth **(A)**, day of starting fortification **(B)** use of antibiotics before start of fortification **(C)** as well as SGA/AGA status at birth **(D)** and their respective association to risk of infection after start of fortification. AB: Antibiotics, AGA: birthweight appropriate for gestational age, SGA: birthweight small for gestational age. Shown as model predicted means with corresponding standard error. *: P < 0.05.

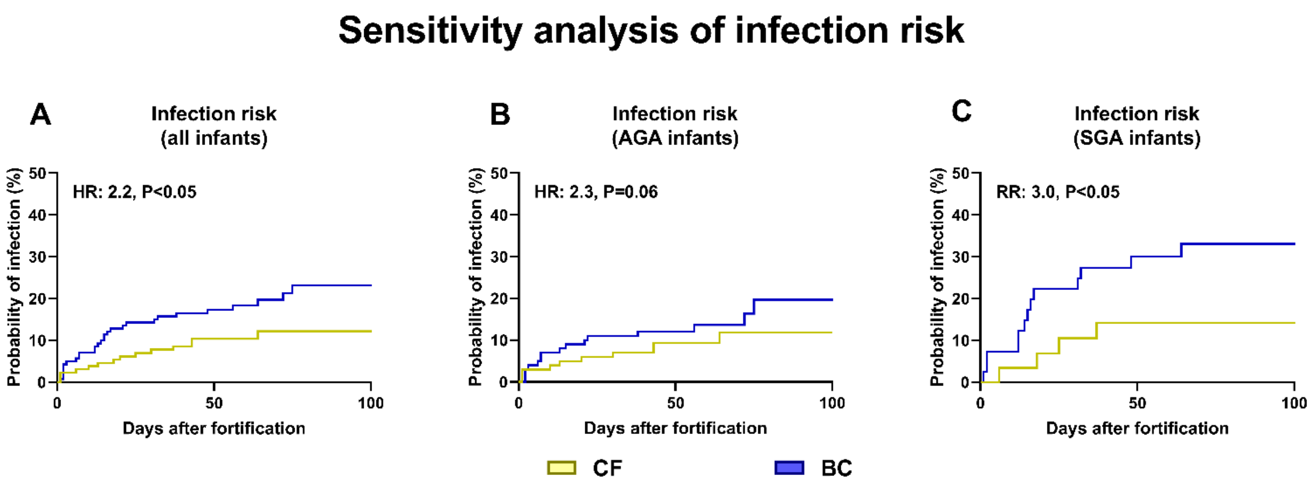


**Supplementary Figure S2**: Risk of infection after start of fortification in infants fortified with bovine colostrum (BC) or conventional fortifier (CF). Sensitivity analysis including only infection cases where plasma levels of C reactive protein were higher than 10 ug/mL and/or blood cultures were positive. Shown for all infants (**A,** n=229) or stratified by birthweight appropriate for gestational age (AGA, **B**) or sSmall for gestational age (SAG, **C**). Shown as Kaplan Meyer curves with results of corresponding Cox proportional hazard models shown as text.

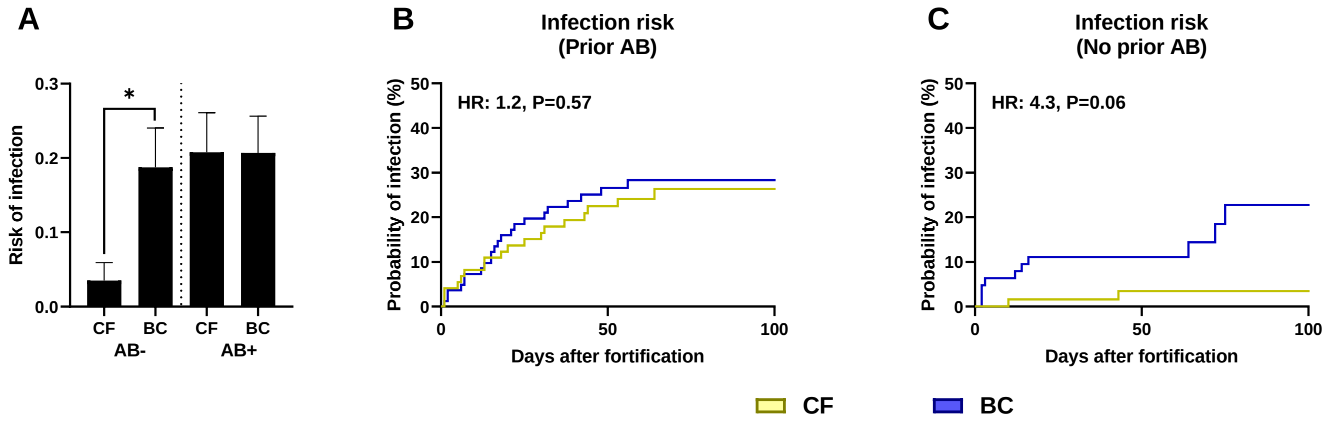


**Supplementary Figure S3**: Incidence of infection after start of fortification in infants fortified with bovine colostrum (BC) or conventional fortifier (CF), stratified by antibiotics (AB) treatment before start of fortification, shown as mean with corresponding standard error, *: P < 0.05. (**A**). Infection risk in infants that did or did not receive AB treatment before start of fortification. Shown as Kaplan Meyer curves with results of corresponding Cox proportional hazard models shown as text (**B, C**). *: P < 0.05.

**Supplementary Table S1: List of intravenous antibiotics used.**

| **Antibiotic** |
|---|
| Ampicillin* |
| Benzylpenicillin |
| Dicloxacillin |
| Cefotaxim |
| Cefuroxim |
| Gentamycin* |
| Meropenem |
| Metronidazol |
| Piperacil/Tazobacam |
| Vancomycin |

*or similar derivative of.

**Supplementary Table 2: Results of clinical blood samples and cultures taken during infection episodes.**

| | All infants | | | AGA | | | SGA | | |
|---|---|---|---|---|---|---|---|---|---|
| | CF n=20 | BC n=32 | P | CF n=16 | BC n=17 | P | CF n=4 | BC n=15 | P |
| **Blood cultures§** | | | | | | | | | |
| Blood culture taken | 45% (9/20) | 53% (17/32) | NS | 43% (7/16) | 65% (11/17) | NS | 50% (2/4) | 40% (6/15) | NS |
| Growth of pathogen | 56% (5/9) | 71% (12/17) | NS | 57% (4/7) | 64% (7/11) | NS | 50 % (1/2) | 83% (5/6) | NS |
| **CRP measurements#** | | | | | | | | | |
| CRP value measured | 75% (15/20) | 88% (28/32) | NS | 75% (12/16) | 95% (16/17) | NS | 75% (3/4) | 80% (12/15) | NS |
| CRP above 10 ug/mL | 80% (12/15) | 86% (24/28) | NS | 75% (9/12) | 75% (12/16) | NS | 100% (3/3) | 100% (12/12) | NS |
| Mean CRP value, ug/mL | 60 (±41) | 76 (±87) | NS | 63 (±42) | 62 (±90) | NS | 49 (±33) | 96 (±81) | NS |
| **Blood gas measurements#** | | | | | | | | | |
| Blood gas measured | 65% (13/20) | 81% (26/32) | NS | 81% (13/16) | 82% (14/17) | NS | 0% (0/4) | 80% (12/15) | - |
| Mean pH value | 7.27 (±0.08) | 7.23 (±0.12) | NS | 7.27 (±0.08) | 7.21 (±016) | NS | - | 7.25 (±0.05) | - |
| Mean lactate value, mmol/L | 1.2 (±0.5) | 1.4 (±0.7) | NS | 1.2 (±0.5) | 1.6 (±1.0) | NS | - | 1.2 (±0.3) | - |

Blood culture and biochemical results in very preterm infants fortified with either conventional fortifier (CF) or bovine colostrum (BC) for all episodes of infection, defined as antibiotic treatment for 5 or more days. Results from blood cultures taken 3 days before and up to 14 days after start of antibiotic treatment. Results from clinical blood samples collected from 3 days before and up to 7 days after start of antibiotic treatment. Shown for all infants and stratified for birth weight status. AGA: Birthweight appropriate-for-gestational age, CRP: C-reactive protein, SGA: Small-for-gestational age, NS: Not significant. Shown as percentages or means with corresponding standard error.

### Supplementary Table S3: Plasma cytokine levels in infants without infection

| | | All infants | | | AGA | | | SGA | | |
|---|---|---|---|---|---|---|---|---|---|---|
| | Day | CF | BC | P | CF | BC | P | CF | BC | P |
| Eotaxin | 0 | 327 (155) | 358 (166) | NS | 325 (158) | 369 (176) | NS | 338 (148) | 315 (117) | NS |
| | 7 | 381 (193) | 420 (197) | NS | 373 (186) | 419 (198) | NS | 411 (222) | 425 (199) | NS |
| | 14 | 334 (184) | 372 (161) | NS | 326 (193) | 378 (158) | 0.06 | 365 (143) | 339 (182) | NS |
| Eotaxin-3 | 0 | 65.9 (91.4) | 101 (235) | NS | 70.2 (100) | 68.4 (95.9) | NS | 48.3 (39.2) | 231 (481) | NS |
| | 7 | 62.7 (143) | 67.9 (129) | NS | 63.9 (157) | 71.1 (141) | NS | 58.1 (72.8) | 54.9 (63.4) | NS |
| | 14 | 63.5 (85.0) | 65.9 (103) | NS | 65.2 (87.0) | 58.0 (64.9) | NS | 56.5 (79.0) | 112 (222) | NS |
| GM-CSF | 0 | 1.61 (4.50) | 0.85 (1.08) | NS | 1.52 (4.34) | 0.93 (1.18) | NS | 1.98 (5.24) | 0.5 (0.32) | NS |
| | 7 | 1.80 (4.20) | 1.38 (3.39) | NS | 1.43 (2.39) | 1.48 (3.71) | NS | 3.31 (8.14) | 0.99 (1.47) | NS |
| | 14 | 1.44 (2.92) | 1.32 (3.47) | NS | 1.34 (2.96) | 1.46 (3.74) | NS | 1.87 (2.77) | 0.51 (0.61) | <0.05 |
| IFN-γ | 0 | 21.0 (70.5) | 25.1 (74.9) | NS | 24.3 (78.2) | 21.8 (70.5) | NS | 7.18 (5.55) | 38.8 (92.4) | NS |
| | 7 | 13.1 (34.6) | 12.1 (22.7) | NS | 9.48 (12.2) | 13.2 (25.1) | NS | 27.0 (71.7) | 7.54 (5.13) | NS |
| | 14 | 20.3 (43.6) | 24.4 (91.8) | NS | 18.9 (40.0) | 21.9 (91.2) | NS | 25.5 (57) | 39.1 (98.5) | NS |
| IL-10 | 0 | 13.3 (44.6) | 15.1 (39.5) | NS | 16.1 (49.3) | 16.8 (42.9) | NS | 1.67 (2.08) | 8.37 (20.1) | NS |
| | 7 | 17.9 (60.6) | 127 (984) | <0.01 | 21.4 (67.5) | 156 (1099) | <0.05 | 4.79 (12.4) | 8.30 (17.2) | <0.001 |
| | 14 | 13.0 (38.2) | 15.1 (44.2) | 0.09 | 15.4 (42.2) | 12.3 (26.7) | NS | 3.50 (9.04) | 31.7 (98.2) | <0.05 |
| IL-12p40 | 0 | 266 (128) | 285 (154) | NS | 272 (129) | 281 (138) | NS | 241 (121) | 298 (213) | NS |
| | 7 | 347 (135) | 337 (137) | NS | 356 (139) | 336 (126) | NS | 311 (111) | 341 (179) | NS |
| | 14 | 415 (156) | 405 (168) | NS | 415 (155) | 406 (165) | NS | 415 (166) | 402 (190) | NS |
| IL-12p70 | 0 | 0.62 (1.99) | 0.82 (3.40) | NS | 0.70 (2.21) | 0.92 (3.78) | NS | 0.27 (0.26) | 0.41 (0.62) | NS |
| | 7 | 0.30 (0.34) | 0.28 (0.26) | NS | 0.31 (0.37) | 0.28 (0.27) | NS | 0.23 (0.15) | 0.29 (0.22) | NS |
| | 14 | 0.29 (0.43) | 0.27 (0.34) | NS | 0.31 (0.48) | 0.24 (0.27) | NS | 0.20 (0.15) | 0.49 (0.57) | NS |
| IL-13 | 0 | 16.1 (31.1) | 14.9 (27.7) | NS | 17.6 (34.3) | 13.9 (23.6) | NS | 9.89 (7.77) | 18.8 (41.0) | NS |
| | 7 | 12.4 (20.0) | 18.4 (33.0) | 0.07 | 13.2 (21.8) | 17.9 (29.8) | NS | 9.39 (10.4) | 20.0 (45.0) | 0.07 |
| | 14 | 15.1 (36.6) | 23.2 (77.7) | NS | 16.3 (39.9) | 18.9 (66.1) | NS | 10.2 (17.7) | 48.2 (128) | <0.05 |
| IL-15 | 0 | 3.82 (4.43) | 4.44 (5.50) | NS | 3.53 (3.69) | 4.61 (5.83) | NS | 5.05 (6.70) | 3.74 (4.01) | NS |
| | 7 | 4.02 (6.17) | 2.47 (3.41) | <0.05 | 3.66 (5.58) | 2.54 (3.66) | NS | 5.48 (8.18) | 2.19 (2.15) | <0.05 |
| | 14 | 5.03 (9.70) | 3.82 (6.27) | NS | 3.57 (7.19) | 4.07 (6.71) | NS | 10.9 (15.2) | 2.35 (2.16) | NS |
| IL-16 | 0 | 419 (998) | 317 (353) | NS | 335 (503) | 300 (213.3) | NS | 766 (2032) | 389 (681) | NS |
| | 7 | 406 (1089) | 238 (123) | NS | 447 (1213) | 243 (131) | NS | 242 (93.0) | 221 (84.6) | NS |
| | 14 | 238 (194) | 216 (80.0) | NS | 240 (212) | 218 (83.4) | NS | 229 (101) | 200 (56.0) | NS |
| IL-17 | 0 | 10.2 (15.6) | 9.83 (11.7) | NS | 10.6 (16.9) | 10.0 (10.5) | NS | 8.59 (8.29) | 9.29 (16.3) | NS |
| | 7 | 14.4 (14.3) | 12.1 (11.6) | NS | 14.7 (15.4) | 13.3 (12.4) | NS | 13.4 (8.82) | 7.11 (4.87) | <0.05 |
| | 14 | 17.4 (12.2) | 15.5 (10.5) | NS | 17.2 (12.2) | 14.8 (8.65) | NS | 17.8 (12.3) | 19.0 (18.2) | NS |
| IL-1α | 0 | 43.3 (54.9) | 38.2 (37.5) | NS | 44.1 (59.3) | 38.7 (36.8) | NS | 40.1 (31.3) | 36.4 (41.9) | NS |
| | 7 | 35.9 (43.3) | 32.4 (40.2) | <0.05 | 36.2 (42.5) | 32.6 (40.2) | 0.05 | 34.4 (47.9) | 31.7 (41.7) | NS |

| | | | | | | | | | | |
|---|---|---|---|---|---|---|---|---|---|---|
| | 14 | 33.2 (66.6) | 42.9 (83.2) | NS | 36.1 (73.9) | 46.9 (89.4) | NS | 21.8 (15.6) | 19.7 (14.9) | <0.05 |
| IL-1β | 0 | 1.42 (2.82) | 0.84 (1.27) | NS | 1.12 (2.25) | 0.83 (1.22) | NS | 2.71 (4.35) | 0.89 (1.50) | NS |
| | 7 | 0.89 (2.48) | 0.72 (1.66) | NS | 0.89 (2.65) | 0.73 (1.71) | NS | 0.93 (1.74) | 0.7 (1.51) | NS |
| | 14 | 0.40 (0.45) | 0.35 (0.40) | 0.05 | 0.35 (0.33) | 0.35 (0.33) | NS | 0.56 (0.76) | 0.37 (0.69) | <0.01 |
| IL-2 | 0 | 0.67 (0.42) | 0.74 (0.70) | NS | 0.67 (0.43) | 0.74 (0.73) | NS | 0.69 (0.34) | 0.76 (0.57) | NS |
| | 7 | 0.73 (0.74) | 0.83 (0.92) | NS | 0.77 (0.81) | 0.80 (0.95) | NS | 0.54 (0.27) | 0.95 (0.80) | NS |
| | 14 | 0.71 (0.54) | 0.71 (0.78) | NS | 0.74 (0.58) | 0.70 (0.80) | NS | 0.58 (0.33) | 0.73 (0.69) | NS |
| IL-4 | 0 | 0.57 (1.17) | 0.57 (1.27) | NS | 0.62 (1.28) | 0.62 (1.41) | NS | 0.35 (0.43) | 0.34 (0.36) | NS |
| | 7 | 0.67 (1.80) | 0.70 (1.38) | NS | 0.77 (1.99) | 0.77 (1.52) | NS | 0.32 (0.62) | 0.44 (0.43) | <0.05 |
| | 14 | 0.78 (2.90) | 0.54 (0.79) | NS | 0.87 (3.22) | 0.54 (0.83) | NS | 0.44 (0.74) | 0.53 (0.53) | NS |
| IL-5 | 0 | 3.03 (6.40) | 2.57 (2.11) | NS | 3.00 (7.03) | 2.64 (2.18) | 0.07 | 3.14 (2.47) | 2.31 (1.84) | NS |
| | 7 | 2.72 (3.78) | 1.88 (1.48) | 0.05 | 2.41 (2.95) | 1.92 (1.56) | NS | 3.99 (6.06) | 1.71 (1.17) | <0.05 |
| | 14 | 2.19 (2.80) | 1.85 (1.52) | NS | 1.88 (1.37) | 1.87 (1.59) | NS | 3.43 (5.58) | 1.72 (1.11) | NS |
| IL-6 | 0 | 8.15 (10.3) | 18.1 (60.6) | NS | 8.30 (11.0) | 18.2 (65.5) | NS | 7.51 (7.36) | 17.6 (36.1) | NS |
| | 7 | 5.34 (6.80) | 4.89 (7.28) | NS | 5.42 (6.93) | 4.52 (7.23) | NS | 5.03 (6.47) | 6.40 (7.53) | <0.05 |
| | 14 | 4.89 (7.07) | 4.12 (11.1) | NS | 5.33 (7.60) | 4.16 (11.9) | NS | 3.12 (3.99) | 3.90 (4.12) | NS |
| IL-7 | 0 | 5.78 (6.62) | 4.91 (3.87) | NS | 5.32 (6.08) | 5.05 (3.90) | NS | 7.66 (8.47) | 4.31 (3.82) | NS |
| | 7 | 4.92 (4.99) | 4.70 (3.47) | NS | 4.47 (3.68) | 4.91 (3.69) | NS | 6.73 (8.37) | 3.87 (2.28) | NS |
| | 14 | 4.13 (4.13) | 4.95 (3.77) | 0.06 | 3.77 (3.42) | 5.20 (3.90) | <0.05 | 5.54 (6.16) | 3.44 (2.56) | NS |
| IL-8 | 0 | 482 (861) | 431 (570) | NS | 365 (469) | 402 (553) | NS | 961 (1649) | 547 (642) | NS |
| | 7 | 432 (716) | 414 (668) | NS | 413 (741) | 423 (698) | NS | 504 (622) | 378 (546) | NS |
| | 14 | 421 (689) | 324 (510) | NS | 415 (676) | 293 (470) | 0.09 | 447 (761) | 510 (699) | NS |
| IP10 | 0 | 358 (568) | 413 (634) | NS | 380 (626) | 369 (598) | NS | 270 (171) | 589 (757) | NS |
| | 7 | 266 (227) | 284 (185) | NS | 248 (201) | 296 (191) | NS | 336 (303) | 233 (153) | NS |
| | 14 | 309 (793) | 252 (207) | NS | 263 (657) | 246 (187) | <0.05 | 497 (1203) | 289 (309) | NS |
| MCP1 | 0 | 215 (140) | 309 (482) | NS | 206 (138) | 273 (435) | NS | 253 (145) | 453 (632) | NS |
| | 7 | 234 (102) | 238 (134) | NS | 228 (104) | 233 (134) | NS | 258 (93) | 257 (133) | NS |
| | 14 | 225 (92.6) | 238 (137) | NS | 214 (87.0) | 245 (138) | NS | 269 (103) | 202 (130) | NS |
| MCP4 | 0 | 115 (91.6) | 115 (59.9) | NS | 98.3 (56.2) | 109 (58.8) | 0.07 | 183 (159) | 139 (60.1) | NS |
| | 7 | 105 (69.6) | 129 (83.4) | 0.07 | 99.7 (71.0) | 119 (78.3) | NS | 125 (61.7) | 168 (93.9) | NS |
| | 14 | 89.0 (54.2) | 106 (59.1) | <0.05 | 84.8 (48.7) | 105 (60.9) | 0.06 | 104 (71.8) | 115 (48.3) | NS |
| MDC | 0 | 992 (593) | 936 (353) | NS | 1012 (636) | 923 (321) | NS | 908 (368) | 992 (469) | NS |
| | 7 | 1024 (581) | 1075 (430) | NS | 1031 (536) | 1076 (439) | NS | 999 (745) | 1068 (405) | <0.01 |
| | 14 | 985 (461) | 1046 (464) | NS | 956 (382) | 1051 (440) | NS | 1102 (697) | 1021 (613) | NS |
| MIP1α | 0 | 59.5 (67.4) | 60.9 (70.2) | NS | 59.4 (67.6) | 53.5 (61.0) | NS | 59.8 (68.6) | 90.3 (95.8) | NS |
| | 7 | 48.5 (63.8) | 46.9 (56.1) | NS | 49.0 (61.1) | 45.4 (51.5) | NS | 46.5 (74.7) | 53.0 (73.8) | NS |
| | 14 | 63.2 (80.9) | 67.1 (73.8) | NS | 62.8 (73.9) | 66.5 (71.6) | NS | 64.6 (107) | 70.6 (89.5) | NS |
| MIP1β | 0 | 194 (139) | 249 (294) | 0.06 | 194 (149) | 221 (173) | NS | 195 (90.4) | 363 (561) | NS |
| | 7 | 166 (92.7) | 169 (88.6) | NS | 164 (88.2) | 171 (88.2) | NS | 176 (110) | 165 (93.2) | NS |

| | | | | | | | | | | |
|---|---|---|---|---|---|---|---|---|---|---|
| | 14 | 152 (77.9) | 157 (74.3) | NS | 147 (78.3) | 161 (74.7) | NS | 171 (75.6) | 135 (70.8) | NS |
| TARC | 0 | 627 (690) | 620 (606) | NS | 650 (714) | 645 (621) | NS | 532 (589) | 522 (551) | NS |
| | 7 | 677 (595) | 851 (785) | NS | 690 (627) | 828 (815) | NS | 630 (468) | 946 (669) | NS |
| | 14 | 416 (360) | 558 (540) | 0.08 | 423 (379) | 597 (566) | <0.05 | 386 (279) | 334 (273) | NS |
| TNF-α | 0 | 3.41 (2.25) | 3.95 (3.36) | NS | 3.32 (1.98) | 3.74 (2.53) | NS | 3.80 (3.18) | 4.84 (5.64) | NS |
| | 7 | 3.03 (1.82) | 3.01 (1.62) | NS | 2.85 (1.30) | 3.15 (1.74) | NS | 3.72 (3.06) | 2.47 (0.84) | NS |
| | 14 | 2.98 (1.77) | 2.99 (1.53) | NS | 2.97 (1.87) | 3.03 (1.58) | NS | 3.01 (1.37) | 2.79 (1.24) | NS |
| TNF-β | 0 | 1.12 (2.60) | 1.77 (4.33) | NS | 1.20 (2.86) | 1.82 (4.69) | NS | 0.81 (0.87) | 1.55 (2.49) | NS |
| | 7 | 0.94 (1.45) | 1.25 (2.90) | NS | 0.81 (1.08) | 1.45 (3.21) | NS | 1.47 (2.42) | 0.46 (0.24) | NS |
| | 14 | 1.67 (3.63) | 1.27 (2.19) | NS | 1.72 (3.70) | 1.26 (2.29) | NS | 1.46 (3.46) | 1.34 (1.58) | NS |
| VEGF | 0 | 302 (304) | 310 (289) | NS | 322 (325) | 337 (307) | NS | 217 (179) | 196 (158) | NS |
| | 7 | 275 (284) | 271 (234) | NS | 276 (303) | 249 (234) | NS | 274 (196) | 361 (218) | NS |
| | 14 | 177 (217) | 183 (143) | NS | 174 (208) | 191 (147) | NS | 187 (254) | 134 (112) | NS |

Plasma cytokine levels in very preterm infants fortified with either conventional fortifier (CF) or bovine colostrum (BC) with no suspicion of infection. Shown for all infants and stratified for birth weight status. AGA: Birthweight appropriate-for-gestational age, SGA: Small-for-gestational age, NS: Not significant, IL: Interleukin, GM-CSF: Granulocyte-macrophage colony stimulating factor, IFN: Interferon, IP10: Interferon gamma-induced protein 10, MCP: Monocyte chemoattractant protein, MDC: Macrophage-derived chemokine, MIP: Macrophage inflammatory protein, TARC: Thymus and activation regulated *chemokine*, TNF: Tumor necrosis factor, VEGF: Vascular endothelial growth factor. Data shown as means with corresponding standard deviation.
